# An investigation of the impact of index case screening on commonly reported epidemiological estimates in tuberculosis (TB) household contact study

**DOI:** 10.1101/2023.03.10.23287100

**Authors:** Yangmingqiu Dou, Tenglong Li

## Abstract

Tuberculosis, a chronic infectious disease caused by Mycobacterium tuberculosis (Mtb), remains as one of the biggest threats to public health worldwide. The purpose of this study is to investigate the impact of different screening criteria of the index cases on epidemiological estimates in the tuberculosis (TB) household contact study. Six different screening criteria were designed based on whether the index cases were adults and lived in the crowded environment, as well as their sputum smear and culture test results. Logistic regression was performed to determine the risk factors of TB. We found that gender, HIV-infection, smoking, malnutrition and diabetes were associated with Mtb infection. We also found significant differences of epidemiological estimates among observation groups screened by different criteria. The number of infections were relatively higher in the studies where index cases were screened as positive sputum smear and living in the crowded environment, indicating that index cases with positive sputum smear and live with more than two contacts in one room were likely to be more infectious in the household. Therefore, it is suggested that screening and treatment should be strengthened for those designs so that transmission of Mtb infection in the household can be reduced.

## 1 Introduction

As one of the oldest infectious diseases, human still suffers from tuberculosis, with an estimated 9.9 million new TB cases and 1.5 million TB deaths worldwide in 2020, according to the World Health Organization’s Global TB Report 2021 [1]. Tuberculosis (TB) is a chronic infectious disease caused by Mycobacterium tuberculosis (Mtb), remaining one of the most important threats to human health all around the world [2]. Mtb infection is primarily via person-to-person respiratory transmission, and those who progress to TB disease can have symptoms such as low fever, fatigue and cough. About one third of the world’s population is infected with Mycobacterium tuberculosis, with 9 million new cases and 2 million deaths from the disease each year [3]. TB caused by Mtb infection can seriously affect human life and global health.

In 2020, there were an estimated 9.9 million new TB cases worldwide, with an incidence rate of 127/100 000 [1]. Both the estimated incidence and incidence rate have shown a downward trend in recent years. Most high-burden countries have rates of TB between 150 and 400 per 100 000, with a few countries, such as Central Africa, North Korea, the Philippines and South Africa having TB rates above 500 per 100 000 [4]. The top eight countries with the highest number of cases were India (2.6 million, 26%), China (0.84 million, 8.5%), Indonesia (0.82, 8.4%), Philippines (0.59 million, 6.0%), Pakistan (0.57 million, 5.8%), Nigeria (0.45 million, 4.6%) and Bangladesh (0.36 million, 3.6%) and South Africa (0.33 million, 3.3%) [1]. Drug-resistant TB also remains a serious global public health crisis. The resistance to two most effective first-line drugs which are isoniazid and rifampicin has raised public concern. Multidrug-resistant tuberculosis (MDR-TB) is defined as resistance to both drugs (and maybe others as well). From 2019 to 2020, the total number of drug-resistant tuberculosis decreased by 22%, and 157903 new drug-resistant tuberculosis cases were detected. Meanwhile, the total number of notifications of TB cases dropped by 18% between 2019 and 2020 [5].

COVID-19 emerged as the leading cause of death among infectious diseases in 2020, reversing a 15-year decline in TB deaths and increasing by 5.6% from 2005, mainly due to disruptions in TB treatment services caused by the pandemic [6]. Since the end of 2019, the COVID-19 pandemic has been raging around the world, the long and widespread pandemic has caused global TB control to set back by 5–8 years, reversing the progress made in recent years in improving TB diagnosis and treatment. First, the number of registered new cases of TB fell from 7.1 million to 5.8 million in 2020 compared with 2019 [2], as the pandemic has made a large number of TB patients not be diagnosed and treated in time, which significantly reduced number of visits and access to medical care for TB patients. Second, the COVID-19 pandemic led to an increase in the number of TB deaths from 1.41 million in 2019 to 1.49 million in 2020 due to the barriers to access for patients, which is the first increase in nearly 15 years [7]. Third, human and financial resources of many countries for fighting TB have been diverted to the response to the COVID-19 pandemic, resulting in a sharp reduction in the global investment in TB treatment and prevention [6]. In the face of the severe challenges posed by COVID-19, WHO calls on all countries to pool their resources for TB prevention and control and accelerate the pace of ending TB.

One of the main goals of global TB control is to reduce the TB transmission. In about 5% of people who have latent TB infection (LTBI) will rapidly develop into TB cases in the first several years after infection [8]. Deepening the understanding of risk factors with regard to the transmission and disease progression of TB is important for TB control and prevention. The risk of infection after the exposure of TB is mainly determined by environmental factors associated with infectees and the infectiousness of infectors [9]. For example, sleep in the same room with a TB case, the number of windows, and housing crowdedness may increase the risk of TB transmission. The microscopic examination of sputum smear for the presence of Mycobacterium tuberculosis is used to diagnose TB all around the world [10]. The smear positive pulmonary TB patients are likely to be more infectious [11]. The risk factors of the progression to TB disease are more likely due to the infectees themselves, such as Mtb and HIV co-infection, smoking, alcoholism, malnutrition and diabetes

The primary objective of TB household contact study is to investigate the dynamics of TB transmission in the context of household contact [12]. The index case is typically defined as a smear-positive adult TB case who has close household contacts [13]. Household contacts are typically defined as individuals who had lived in the same house with the index case for a cumulative of at least 7 days within the past three months before the index case was diagnosed [14]. In a Brazilian household contact study design, households will be enrolled if the TB case in the household satisfy the following criteria, 1) adults, 2) positive tuberculosis sputum smear, 3) positive sputum culture, 4) living with three or more household contacts [12–13]. The essence of TB household contact study is to select all members in the household who have been in close contact with the TB index case and collect their epidemiological data for TB research [15–16]. The design of household contact study is appropriate for researching TB transmission from the perspective of epidemiology as there exists higher risks for household contacts of TB cases to infect TB due to intimate contacts [17].

The design of TB household contact studies differs from that of influenza. In influenza household contact studies, selected cohort of households are enrolled before the influenza season begins, and all of these households are followed up after the finish of the influenza season to collect their data to determine the infection status of household members. However, in the TB household contact study, only households with the index cases are enrolled to study TB transmission dynamics, and households without TB cases are ignored, which may bring the selection bias.

In this study, simulated data is used to explore TB transmission dynamics in a pre-designed population. The transmission dynamics of our simulation consists of two transmission forces, namely community transmission and household transmission, and two separate models are established for these two different transmission forces. The features of the general population as well as households are inputted, and then TB transmission paths are simulated in each household. There are 8 covariates in total and they are divided into two groups. The first group is the fixed factors. The proportion of covariates in this group is fixed and does not change with gender and age. Among all the households, 60% of them are set as crowded, 70% of the population are set as adults, and 50% of them are women. Each of the four communities accounts for 25 per cent of the population. The covariates in the second group, including smoking, malnutrition, HIV and diabetes, do depend on age and gender. We need to interact these four covariates with gender and whether they are adults respectively to form four groups with different probability, so as to achieve the goal of stratification of the population. The input for these unfixed covariates should be a matrix of 4*4, columns for (1)adult male (2)adult female (3)non-adult male (4)non-adult female. In this way, we can more accurately simulate the prevalence of these risk factors.

Since only households with the index cases are enrolled in TB household contact studies, then the screening criteria or eligibility of the index cases is an important issue. As mentioned above, the index case screening criteria in most TB household contact studies are as follows: 1-adult; 2-positive sputum smear test result; 3-positive culture test results; 4-living with three or more household contacts. However, the above index case screening criteria may lead to bias in some epidemiological estimates such as second attack rate, annual TB infection rate and prevalence of TB cases. The research goal of this paper is to explore the impact of different index case screening criteria on common epidemiological estimates via a simulation study.

## 2 Methods

### 2.1 Epidemiological estimates in TB research

Odds ratio (OR) is a measure that quantifies the relationship between the exposure status to a risk factor and infection status. It is calculated as the ratio of the number of people exposed and with disease to the number of people exposed but without disease divided by the ratio of the number of people unexposed but with disease to the number of people unexposed and without disease [18]. If the OR value is equal to 1, it indicates that the factor has no effect on the occurrence of the disease. If the OR value is greater than 1, it indicates that the risk factor is harmful. If the OR value is less than 1, it indicates that the factor is not harmful (but beneficial).

#### 2.1.2 Second attack rate

The second attack rate (SAR) refers to the percentage of household contacts who develop TB disease during the observation period after the first case of infectious disease in the household. In this study, it is calculated as dividing the number of new TB cases resulting from contacts with (and share the same strain with) the primary TB cases by the total number of contacts. What should be emphasized is that the total number of contacts in the denominator must be calculated by removing the primary TB case from the total number of the household members [18]. The second attack rate can reflect the household transmission force, and is used to evaluate the effect of epidemic prevention measures and impact of potential risk factors.

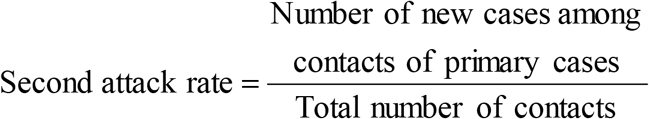

#### 2.1.3 Prevalence of TB cases

The prevalence of TB cases is the proportion of people in a population who have TB disease at a particular point in time or during a particular period of time. TB prevalence is measured in a population at a given time and includes both new cases and all existing cases, rather than being limited to the number of new cases [18]. The period prevalence of TB is calculated as the number of new and primary cases of TB in a certain population during the observation period divided by the population during the same period. TB prevalence and incidence are often confused. To be specific, TB prevalence is the proportion of people have TB disease at a given time. TB incidence, however, is the proportion of people who develop TB disease during a given period of time.

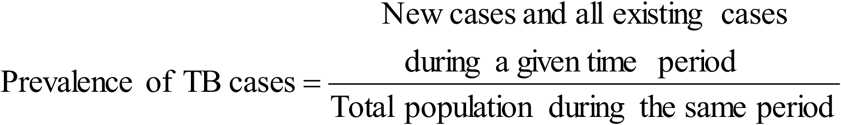

#### 2.1.4 Annual infection rate

The annual TB infection rate is calculated as the number of TB infections in a population divided by the total number of people multiplied by the total observation time. The denominator of the annual infection rate requires to be calculated separately by TB cases and people who are not TB cases. For TB cases, the total number of cases should be multiplied by the corresponding onset time (years since the start of the observation). For others, the population size is multiplied by the total years of observation.

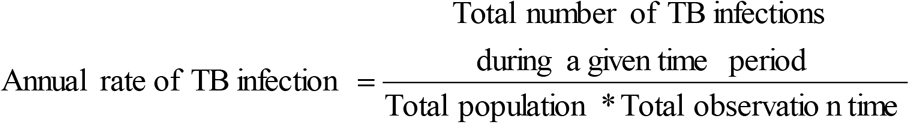

### 2.2 Index case screening

Screening of the TB index case is defined as the systematic identification of TB patients in a target population via a test, examination, or other procedure that can be applied rapidly [19]. The criteria of screening should effectively focus on people who are at high risk of developing TB. Household contacts of TB patients have a higher risk of exposure to pathogenic organisms than the general population. The risk of infection among contacts is related to the infectivity of patients, contact time and individual susceptibility. A systematic review showed that around 5% of household contacts or other close contacts of the index cases had previously undiagnosed infections [11].

The Chinese Ministry of Health piloted a new project to screen suspected TB patients among household contacts of index cases in 2007 with the aim of informing screening strategies that could be implemented in the future [17]. Since there were more positive TB patients being identified in this pilot project than in the routine surveillance of TB disease, more tests among households were rolled out. After each index case was diagnosed, all the household contacts who had shared the same house with the index case for more than two weeks within the past three months before the diagnosis of the index case were referred for screening.

### 2.3 Impact of index case screening

Screening is often seen as proactive case finding and it can target people with or without TB symptoms. Children, the elderly, people with diabetes, people with HIV and other groups whose immune systems have been compromised have higher risk of adverse effects of TB treatment and the risk increases with a delay in diagnosis [19]. Household with the index case is well suited for detecting other TB cases, as household transmission of Mtb is more likely to happen in this case [8]. Studies have shown that smear-positive TB cases are more infectious than smear-negative TB cases, though smear-negative TB cases can also transmit Mtb [10]. Proper index case screening can ensure early detection of TB cases and initiation of the treatment, which can reduce the TB burden and the risk of TB transmission.

### 2.4 Simulation study

Through simulation, we got a dataset with a population of 9937 people consisted of 1927 different households in 4 communities [12]. There were 292 TB cases, 2566 individuals who had latent TB infection (LTBI) and 7079 individuals who were not infected with TB. Based on this simulated dataset, we can test different index case screening criteria for household contact study [20–22]. In order to study the impact of different index case screening criteria on relevant epidemiological estimates, we compared the following sets of index case screening criteria on the TB cases: 1) adults, sputum smear-positive and sputum culture-positive, living in a crowded environment. 2) nonadults with positive sputum smear and sputum culture, living in a crowded environment. 3) adults with positive sputum smear and negative sputum culture, living in a crowded environment. 4) adults with negative sputum smear and positive sputum culture, living in a crowded environment. 5) adults with negative sputum smear and sputum culture, living in a crowded environment. 6) adults with positive sputum smear and sputum culture, living in a non-crowded environment.

All data analyses and calculations were performed using R software. Chi-squared test was used for univariate analysis and logistic regression was used for multivariate analysis of the risk factors. Risk factors with p-values smaller than 0.05 were considered to be statistically significant.

## 3 Results

As Table 1 shows, there were 9937 observations in this study, including a total of 1927 households, 4925 males, 5012 females, 6957 adults and 2980 who were under 18 years old. 5983 people lived in the crowded environment (household that has over 2 inhabitants per inhabitable room). There were 2847 smokers, 1933 malnourished individuals, 697 individuals infected with HIV and 1426 individuals who had diabetes. In total, there were 292 people who were infected with TB, 2566 people who had latent TB infection and 7079 were not infected with TB.

**Table 1:** Descriptive statistics

| Variables | Total<br>(n = 9937) | Non-infection<br>(n = 7079) | Latent TB infection<br>(n = 2566) | TB disease<br>(n = 292) | P-Value |
| --- | --- | --- | --- | --- | --- |
| Crowd, n (%) |  |  |  |  | 0.321 |
| No | 3954 (39.79) | 2832 (40.01) | 1018 (39.67) | 104 (35.62) |  |
| Yes | 5983 (60.21) | 4247 (59.99) | 1548 (60.33) | 188 (64.38) |  |
| Female, n (%) |  |  |  |  | < 0.001 |
| No | 4925 (49.56) | 3403 (48.07) | 1369 (53.35) | 153 (52.4) |  |
| Yes | 5012 (50.44) | 3676 (51.93) | 1197 (46.65) | 139 (47.6) |  |
| Adult, n (%) |  |  |  |  | < 0.001 |
| No | 2980 (29.99) | 2292 (32.38) | 629 (24.51) | 59 (20.21) |  |
| Yes | 6957 (70.01) | 4787 (67.62) | 1937 (75.49) | 233 (79.79) |  |
| Smoking, n (%) |  |  |  |  | < 0.001 |
| No | 7090 (71.35) | 5345 (75.51) | 1563 (60.91) | 182 (62.33) |  |
| Yes | 2847 (28.65) | 1734 (24.49) | 1003 (39.09) | 110 (37.67) |  |
| Malnutrition, n (%) |  |  |  |  | < 0.001 |
| No | 8004 (80.55) | 5857 (82.74) | 1933 (75.33) | 214 (73.29) |  |
| Yes | 1933 (19.45) | 1222 (17.26) | 633 (24.67) | 78 (26.71) |  |
| HIV, n (%) |  |  |  |  | < 0.001 |
| No | 9240 (92.99) | 6639 (93.78) | 2333 (90.92) | 268 (91.78) |  |
| Yes | 697 (7.01) | 440 (6.22) | 233 (9.08) | 24 (8.22) |  |
| Diabetes, n (%) |  |  |  |  | < 0.001 |
| No | 8511 (85.65) | 6163 (87.06) | 2103 (81.96) | 245 (83.9) |  |
| Yes | 1426 (14.35) | 916 (12.94) | 463 (18.04) | 47 (16.1) |  |

From Table 1, we can see that gender is a significant influencing factor for TB infection. The proportion of males who were not infected is 48.07%, who had latent TB infection is 53.35% and who were infected is 52.4%. It can be seen that there exists difference between men and women regarding the proportions of TB disease and infection, and such difference is statistically significant (p<0.05). Living in the crowded environment is not a significant influencing factor for TB infection. 59.99% of the non-infected individuals, 60.33% of the LTBI individuals and 64.38% of the TB cases were living in the crowded environment, and such difference is not statistically significant (p=0.321). Being adult is one of the significant influencing factors of TB infection. 67.62% of the non-infected individuals were adults and 32.38% of the non-infected individuals were minors. Among the TB cases, 79.79% were adults and while only 20.21% of them were minors. Among LTBI individuals, 75.49% were adults while only 24.51% were minors, indicating that there was difference between the groups of adults and minors, and such difference is statistically significant (p<0.05). Smoking is also one of the significant influencing factors for TB infection. Among non-infected individuals, 24.49% of them were smokers and 75.51% of them were non-smokers. Among the TB cases, 37.67% were smokers and 62.33% were non-smokers. This difference is statistically significant (p<0.001). The proportion of smokers among the TB cases is 1.54 times that of non-infected individuals, which suggests that smokers are more likely to develop TB disease than non-smokers. Malnutrition is another significant influencing factor of TB infection. The proportion of malnutrition was 17.26%, 24.67%, 26.71%, respectively, among non-infected individuals, LTBI individuals and TB cases. Therefore, there is difference between malnourished and nourished individuals, and the difference is statistically significant (p<0.001). The proportion of malnutrition in the infected people was 1.55 times higher than that of the uninfected people, suggesting that the possibility of occurrence of tuberculosis in malnourished people was higher than that in nourished people. HIV infection remains one possible factor of TB infection. HIV-infected people accounted for 6.22% among people who were not infected with TB, 9.08% among people with latent TB infection, and 8.22% among people with TB disease. It was evident that there was relationship between the HIV infection status and the TB infection status, and such relationship was statistically significant (p<0.001). Additionally, people with diabetes tended to be correlated with TB infections. The proportion of people with diabetes among the non-infected individuals was 12.94%, and the proportion of people without diabetes was 87.06% among the non-infected individuals. The proportion of people with diabetes among the LTBI individuals was 18.04%, and the proportion of those without diabetes among the LTBI individuals was 81.96%. The proportion of people with diabetes among the TB case was 16.10%, and proportion of those without diabetes among the TB cases was 83.90%. The relationship between diabetes and the TB infection status was statistically significant (p<0.05).

Logistic regression analysis was performed to determine the risk factors of TB infection. Factors associated with TB infection status in the univariate analysis in Table 1 (p<0.05) were reserved for the multivariate analysis. As shown in Table 2, we found gender, smoking, malnutrition, HIV-infection, and diabetes had statistically significant associations with TB infection status. With regard to TB infections: the odds of smokers was 1.84 (95% CI: 1.66∼2.04) times higher than that of non-smokers; the odds of malnourished people was 1.61 (95% CI: 1.45∼1.79) times higher than that of non-malnourished people; the odds of people with HIV was 2.03 (95% CI: 1.86∼2.56) times higher than that of people without HIV; and the odds of people with diabetes was 1.31 (95% CI: 1.16∼1.49) times higher than that of people without diabetes.

**Table 2.** Logistic regression analysis on TB risk factors

| Variables | Estimate | Std. Errors | OR | 95% CI | P-Value |
| --- | --- | --- | --- | --- | --- |
| Female | -0.11 | 0.05 | 0.89 | 0.82~0.98 | 0.02 |
| Adult | 0.09 | 0.06 | 1.1 | 0.99~1.24 | 0.08 |
| smoking | 0.61 | 0.05 | 1.84 | 1.66~2.04 | <0.001 |
| malnutrition | 0.48 | 0.05 | 1.61 | 1.45~1.79 | <0.001 |
| HIV | 0.28 | 0.08 | 2.03 | 1.86~2.56 | <0.001 |
| diabetes | 0.27 | 0.06 | 1.31 | 1.16~1.49 | <0.001 |

**Table 3:** Odds ratio of four variables in six observation groups

| Variables | First group | Second group | Third group | Fourth group | Fifth group | Sixth group |
| --- | --- | --- | --- | --- | --- | --- |
| Smoking | 3.05 | 1.44 | 4.54 | 2.4 | 1.17 | 2.07 |
| Malnutrition | 2.58 | 3.49 | 1.33 | 0.88 | 1.41 | 1.84 |
| HIV | 1.59 | 1.21 | 3.46 | 3.87 | 0.94 | 1.64 |
| Diabetes | 1.41 | 0.73 | 1.45 | 1.75 | 1.21 | 1.96 |

Epidemiological estimates in the six different designs of screening criteria of the index cases were obtained. The first observation group was selected using the common index case screening criteria, i.e., individuals who were adults, tested positive for sputum smear and culture and lived in the crowded environment. As a result, 101 index cases were selected. All family members in their households were also selected, and there were a total of 578 people in this group, including 279 uninfected, 183 latently infected and 116 TB cases. In this group, variables which were significantly associated with TB infection were smoking (OR=3.05, 95% CI: 2.09∼4.49), malnutrition (OR=2.58, 95% CI: 1.69∼3.99). The second attack rate of TB in this group was 0.428, the annual infection rate was 0.056, the prevalence of TB cases was 0.201, and the prevalence of latent TB infection was 0.317. In the second observation group, the index cases were selected as those who were not adults, tested positive for sputum smear and culture and lived in the crowded environment. This group includes 23 index cases and the size of this group was 134, including 63 uninfected, 41 latent infected and 30 TB cases. Malnutrition had statistically significant association with TB infection (OR=3.49, 95% CI: 1.48∼9.01). The second attack rate in this population was 0.438, the annual TB infection rate was 0.057, the prevalence of TB cases was 0.224, and the prevalence of latent TB infection was 0.306. The third observation group was formed by enrolling the index cases who were adults, living in a crowded environment, tested positive for sputum smear and negative for sputum culture. This group includes 25 index cases and a total of 114 individuals, including 67 uninfected, 44 LTBI and 33 TB cases. Smoking had statistically significant association with TB infection (OR=4.54, 95% CI: 2.05∼10.76). The second attack rate in this population was 0.442, the annual TB infection rate was 0.058, the prevalence of TB cases was 0.229, and the prevalence of latent infection was 0.306. Index cases selected in the fourth observation group were those who were adults, living in a crowded environment, tested negative for sputum smear and positive for sputum culture. This group included 20 index cases and a total number of 109 people, including 57 uninfected, 28 LTBI and 24 TB cases. The second attack rate in this group was 0.359, the annual TB infection rate is 0.047, the prevalence of TB cases was 0.22, and the prevalence of latent TB infection was 0.257. The index cases in the fifth observation group were those who were indeed TB cases but tested negative for sputum smear and culture, adults and living in the crowded environment. This group included only 6 index cases and the total number of people in this group was 33, including 16 uninfected, 7 LTBI and 10 TB cases. The second attack rate in this population was 0.407, the annual TB infection rate was 0.054, the prevalence of TB cases was 0.303, and the prevalence of latent TB infection was 0.212. In the sixth observation group, we selected the index cases as those who were adults and lived in an uncrowded environment, tested positive for sputum smear and culture. This group contained 49 index cases and a total number of 277 people, including 151 uninfected, 74 LTBI and 52 TB cases. Smoking was significantly associated with TB infection (OR=2.07, 95% CI: 1.22∼3.54). The second attack rate in this population was 0.343, the annual TB infection rate was 0.045, the prevalence of TB cases was 0.188, and the prevalence of latent TB infection was 0.267.

## 4 Discussion

This work is mainly divided into two parts. First, relevant risk factors of TB were obtained, based on which constructive suggestions can be provided regarding index case screening. Secondly, epidemiological estimates including the second attack rate, annual TB infection rate, prevalence of TB cases and prevalence of latent TB infection of the study groups screened under different criteria were calculated. As shown in Table 4, epidemiological estimates were relatively different in the groups selected by different TB index case screening criteria, though the variation was not quite large. Relevant conclusions can be drawn regarding the similarities and differences in the epidemiological estimates among the study groups.

**Table 4:** Epidemiological estimates in six observation groups

| Epidemiological estimates | First group | Second group | Third group | Fourth group | Fifth group | Sixth group |
| --- | --- | --- | --- | --- | --- | --- |
| Second attack rate | 0.428 | 0.438 | 0.442 | 0.359 | 0.407 | 0.343 |
| Annual TB infection rate | 0.056 | 0.057 | 0.058 | 0.047 | 0.054 | 0.045 |
| Prevalence of TB cases | 0.201 | 0.224 | 0.229 | 0.22 | 0.303 | 0.188 |
| Prevalence of latent TB infection | 0.317 | 0.306 | 0.306 | 0.257 | 0.212 | 0.267 |

This study showed that HIV infection, diabetes, smoking, malnutrition were significantly associated with TB. The increase of HIV infection has coincided with the increase of TB incidence in the past years. The probability of developing TB in HIV-negative individuals infected with M.tuberculosis (MTB) is only 10%, while the probability of TB in HIV-positive individuals infected with MTB increases to 50%. MTB and HIV can inevitably affect and promote each other, triggering the co-infection of both MTB and HIV. Due to metabolic disorders and weakened immune function, the increase of blood glucose is conducive to the reproduction of MTB [9], rendering diabetes patients more susceptible to pulmonary tuberculosis. Smoking irritates the throat, trachea and lungs, which reduces the defense ability of the respiratory system and endangers people for respiratory infections. Smokers often accompanied by cough and expectoration, which are also the symptoms of pulmonary tuberculosis so that the diagnosis of TB can be delayed. Malnutrition is positively correlated with the risk of TB disease as malnutrition can undermine the immune system, and TB in return can lead to malnutrition due to the changes of the metabolic process.

Previous studies showed that smear positive cases were more infectious and an untreated smear positive TB case could potentially bring about 10 new infections and 2 new TB cases every year [23]. In addition, although the smear negative TB cases are probably less infectious smear positive cases, the TB cases who are smear negative but culture positive are still able to transmit Mtb efficiently [10]. In this study, the first group’s index screening criteria were adults, positive sputum smear and culture, living in the crowded environment. The third group’s index screening criteria were adults, positive sputum smear and negative sputum culture, living in the crowded environment and the fourth group’s index screening criteria were adults, negative sputum smear and positive sputum culture, living in the crowded environment. The epidemiological estimates of second attack rate, annual TB infection rate and prevalence of LTBI in the first and third groups where TB index cases were positive smear are both relatively larger than those in the fourth group which TB index cases were diagnosed as negative sputum smear. By comparing the first, the third and the fourth observation groups, we found that TB index cases that diagnosed with positive sputum smear are more likely to be infectious while index cases with negative sputum smear is also the source of TB transmission within the household. Experimental studies have proved that the infection dose of Mycobacterium tuberculosis can be as low as 1 to 10 bacilli [23]. According to Narasimhan, P., the concentration of bacilli in the sputum of a TB case is positively associated with the infectivity of TB patients, and the number of bacilli in smear negative patients is expected to be less than that in smear positive patients, however the smear negative patients can still transmit Mtb [9].

Compared with the first group, epidemiological estimates in the sixth group were relatively smaller. Recall that the index case screening criteria for the first group were adults, positive sputum smear and sputum culture and lived in the crowded environment while the index case screening criteria for the sixth group were adults, positive sputum smear and sputum culture but lived in an uncrowded environment. Conditional on the TB index cases being smear positive, the index case living in a crowded household may further increase the risk of TB transmission and thus inflate related epidemiological estimates. In this study, a crowded household is defined as household which has over two inhabitants per inhabitable room. Sharing the same room with a smear positive TB case is an important factor associated with the household transmission of TB infection. Household contacts who share the same room with the index case likely have enduring exposures to Mtb than other household contacts, suggesting that they are more likely to be infected.

To sum up, understanding the transmission dynamics of Mtb is central to global TB control and prevention. Early case detection, diagnosis and treatment can reduce the infectiousness of TB cases and shorten the duration of Mtb transmission [23]. The smear positive TB cases are more infectious, so the screening and proper treatment of smear positive index cases, especially those who live in crowded households should be promoted. This is consistent with the current mainstream index screening criteria in household contact study. The index cases with negative sputum smear and positive sputum culture can also effectively transmit Mtb, so screening and diagnosis of the index cases that meet this criteria are also important. Index case screening and preventive treatment are also the keys for preventing the progression to TB disease for high-risk groups. Though, the long latency and measurement error in index case screening may compromise data quality in household contact studies [24]. Through treatment of other morbidities, such as HIV infection, diabetes, malnutrition, along with controlling risk factors such as alcoholism and smoking, the likelihood of infection and progression to TB disease for household can be further reduced.

This study has the limitations that although comparisons were made between distinct observation groups screened under different TB index case screening criteria, the sample sizes of some groups were small and it was unknown that the Mtb transmission characterized in those groups was representative of the whole population. Moreover, there are other risk factors that were not considered in our study, which could undermine the validity of our estimates and therefore robustness analysis maybe warranted [25–28]. In addition, although sputum smear microscopy is mainly used for the diagnosis and treatment scheme of tuberculosis, the wide application of sputum smear microscopy has not resulted in a significant reduction in the incidence rate of TB, which may be partially due to its poor sensitivity [23]. It is recommended that local TB control agencies should adopt new diagnosis methods to improve the sensitivity of TB case detection and diagnosis.

## Data Availability

All data produced in the present study are available upon reasonable request to the authors

